# Participation of Latin America and the Caribbean in vestibular migraine research: a bibliometric study

**DOI:** 10.64898/2026.11.20.26368624

**Authors:** Carlos Arturo Vélez Duncan, Antistio Aníbal Alviz Amador, Dios Isabel Oviedo Tapia, Oscar Daniel Oviedo Tapia

**Author notes:** **Corresponding author:** Carlos Arturo Vélez Duncan, Faculty of Medicine, Universidad de Cartagena, Cartagena, Colombia.

## Abstract

**Background:** How much Latin America and the Caribbean contribute to vestibular migraine research, and whether the region lags further behind in this field than in others, has not been measured.

**Objective:** To compare regional output with that of other world regions and of reference fields, and to contrast the types of studies produced inside and outside the region.

**Methods:** Bibliometric study of PubMed and Scopus records published from 2015 to 2025, reported according to the BIBLIO guideline. Regional participation was compared with seven reference fields in Scopus, with six other World Bank regions and with population; study design was classified with a single prespecified rule.

**Results:** Of 1427 documents, 64 (4.48%; 95% CI 3.53 to 5.69) had at least one affiliation in the region. The region produced 0.09 documents per million inhabitants, compared with 1.04 in North America and 0.66 in Europe and Central Asia (p < 0.001). Its share in Scopus (3.95%) did not differ from its share in migraine (4.31%) or in the whole database (4.77%; p = 0.16), and its specialization index (0.83) did not differ from 1. The distribution of study designs did not differ between regional and non-regional documents (p = 0.88); randomized trials were scarce worldwide.

**Conclusions:** The gap between Latin America and the Caribbean and North America and Europe in vestibular migraine research is wide and similar in magnitude to the gap across the whole database; no additional lag specific to vestibular migraine was detected, although a moderate deficit cannot be ruled out.

## Introduction

Vertigo and dizziness affect about one in five adults over the course of a year^1^. One of their causes is vestibular migraine, which a population-based survey in the United States estimated to affect approximately 2.7% of adults^2^. Consensus diagnostic criteria were issued by the Bárány Society and the International Headache Society in 2012 and updated in 2022^3^.

The evidence for treating it is still limited. Reviews agree that the overall quality of the evidence is low^4^ and that most prophylactic drugs are borrowed from migraine with headache^5^. Much of what is known about antibodies against calcitonin gene-related peptide comes from observational cohorts^6^, and controlled trials are few: a phase III trial of metoprolol was conducted in eight German university hospitals^7^, and the first trial of an antibody of that family was published in 2024^8^.

Health research is unevenly distributed across regions. The mismatch between the research that is needed and the research that is done, known as the 10/90 gap, is well documented^9^. Whether this inequality is uniform, or whether some clinical fields carry an additional regional lag, remains unknown, and that question can only be answered with a comparator.

Bibliometric studies in the area have addressed dizziness in older people^10^, vestibular rehabilitation^11^, benign paroxysmal positional vertigo^12^, calcitonin gene-related peptide in migraine^13^ and the relationship between migraine and anxiety^14^. They describe output by country, but none examines the participation of Latin America and the Caribbean against a baseline, and we did not identify any devoted to vestibular migraine.

We aimed to estimate the participation of Latin America and the Caribbean in the scientific output on vestibular migraine between 2015 and 2025, to compare it with that of other regions, with the region’s participation in reference fields and with its population share, and to contrast the design and orientation of studies produced inside and outside the region. We hypothesized that regional participation in vestibular migraine would be lower than in the reference fields.

## Methods

### Study design

Descriptive and comparative bibliometric study, reported according to the BIBLIO guideline^15^; the selection flow is presented in a diagram based on PRISMA 2020^16^.

Sources and search. PubMed/MEDLINE and Scopus were searched on 12 October 2026. The search combined thirteen expressions in English, Spanish and Portuguese (“vestibular migraine”, “migrainous vertigo”, “migraine-associated vertigo”, among others) in title, abstract and keywords, plus the MeSH descriptor “Vestibular Migraine” in PubMed, for publications from 1 January 2015 to 31 December 2025, without language restriction. The verbatim search strings are given in the supplementary material. A single worldwide corpus was built, of which the regional set is a subset.

### Selection

Records from both databases were matched by PubMed identifier, DOI or, failing those, normalized title and year. Documents outside the period, errata, books and book chapters, and second versions of the same work were excluded. No record was excluded for lacking a DOI, authors or keywords.

### Geographic assignment

Full counting was used: a document counts once for each country with at least one affiliated author^17^. Latin America and the Caribbean comprised the 33 states of the region according to the United Nations, plus Puerto Rico. Country was obtained from the Scopus affiliation index and from each author’s affiliations in PubMed (98.4% of 6243 resolved); in the 849 documents with both sources, the regional classification agreed in all cases. Details and sensitivity analyses are given in the supplementary material.

### Comparators and regions

Using the same period, the same database (Scopus) and the same country rule, regional participation was calculated in seven reference sets: migraine; vertigo, dizziness and vestibular disorders; benign paroxysmal positional vertigo; Ménière’s disease; neuroscience; medicine; and the whole database. For the seven World Bank regions we calculated documents per million inhabitants (mean population 2015 to 2025) and a specialization index: the region’s share in vestibular migraine divided by its share in the whole database. An index of 1 indicates that the region contributes to this field the same as to any other.

### Document classification

A single rule, based on PubMed publication types, Scopus document type and title, classified each document as randomized controlled trial, other trial or protocol, systematic review or meta-analysis, narrative review, case report or series, observational study or other original, or letter, editorial or commentary. Therapeutic orientation was also flagged. Documents naming the condition in the title or author keywords formed a focal subset.

### Statistical analysis

Proportions are presented with 95% confidence intervals (Wilson method). We used Fisher’s exact test to compare shares, the two-sample Poisson test for per-capita rates, the exact binomial test for the specialization index, the Cochran-Armitage test for annual trend, chi-squared with a permutation p-value (9999 resamples) for the distribution of designs, and the Mann-Whitney test for citations. Analyses were performed in Python 3.14 (Python Software Foundation, Wilmington, DE, USA) with pandas 3.0 and SciPy 1.17. The code for retrieval, cleaning, analysis and figures was written with the assistance of a large language model (Claude, Anthropic); the authors reviewed the code and verified the results against the data.

### Ethical considerations

The study analyzed only publicly available bibliographic metadata of scientific publications, with no human participants and no access to personal or clinical data. It qualifies as research without risk under article 11 of Resolution 8430 of 1993 of the Colombian Ministry of Health^18^.

## Results

### Corpus

The searches returned 2416 records (948 from PubMed and 1468 from Scopus). After removing 899 duplicates and excluding 90 documents, the corpus comprised 1427 documents (Figure 1). Country could not be determined in 21 (1.5%). Annual worldwide output in Scopus rose from 83 documents in 2015 to 213 in 2025. Seventy-six countries participated, led by the United States (341 documents; 23.9%), China (238; 16.7%) and Germany (129; 9.0%).

**Figure 1.**
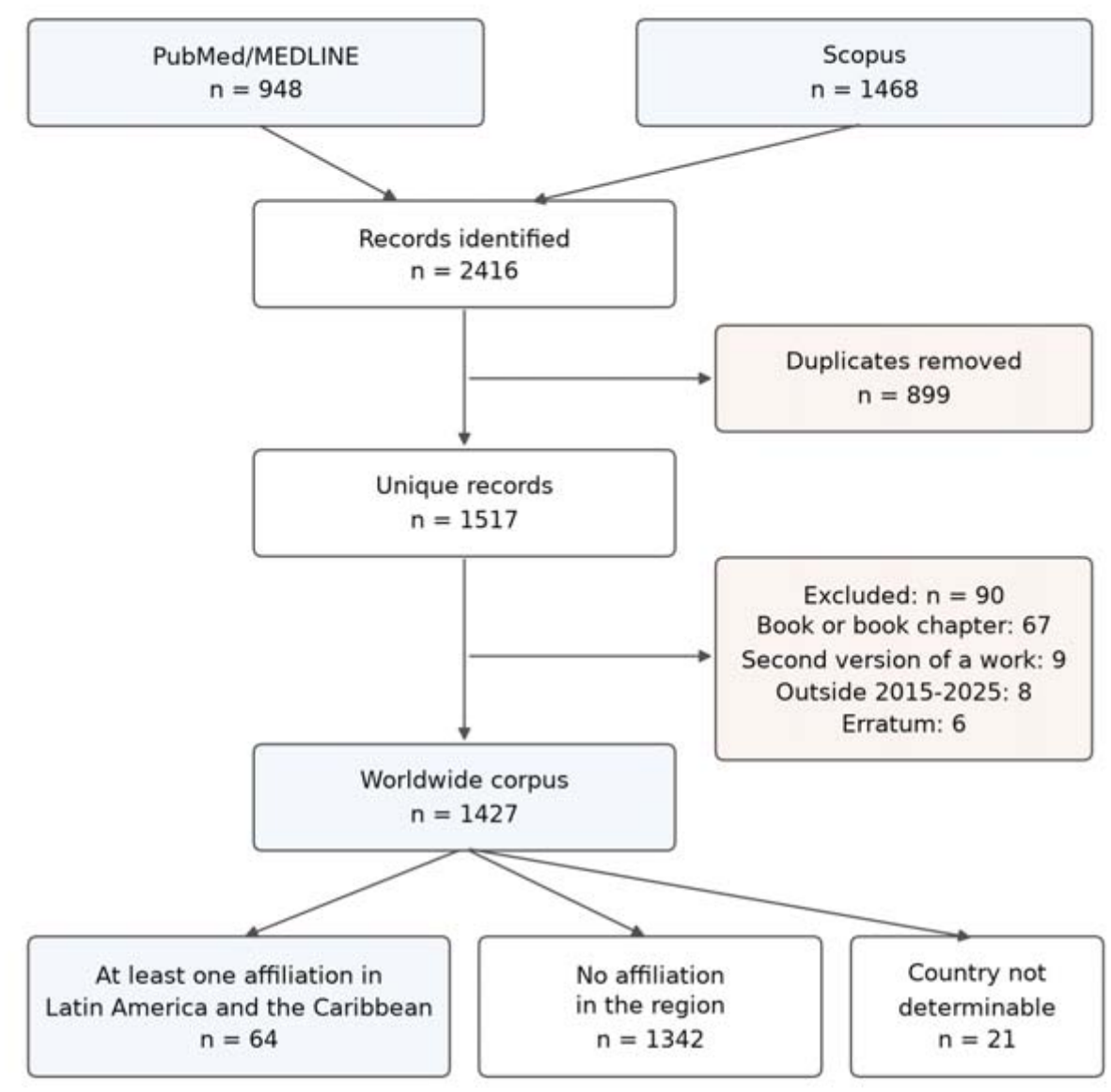
Identification and selection of records.

### Regional participation and comparators

Sixty-four documents had at least one affiliation in the region: 4.48% of the corpus (95% CI 3.53 to 5.69); in the focal subset they were 32 of 763 (4.19%), and in 47 of the 64 the first author had a regional affiliation. In Scopus, regional participation in vestibular migraine was 3.95% (58 of 1468; 95% CI 3.07 to 5.07), similar to that in migraine (4.31%; p = 0.55), vertigo, dizziness and vestibular disorders (3.85%; p = 0.84), neuroscience (4.67%; p = 0.22), medicine (4.73%; p = 0.18) and the whole database (4.77%; p = 0.16); it was higher only than in Ménière’s disease (2.77%; p = 0.03) (Figure 2a). The hypothesis of lower participation was not supported. The region accounted for 8.21% of the world population: it published 58 documents, whereas its population share would correspond to 120 and its share of the whole database to 70. Its participation showed no trend between 2015 and 2025 (p = 0.47), whereas in migraine it increased from 3.48% to 5.44% (p < 0.001) (Figure 2b).

**Figure 2.**
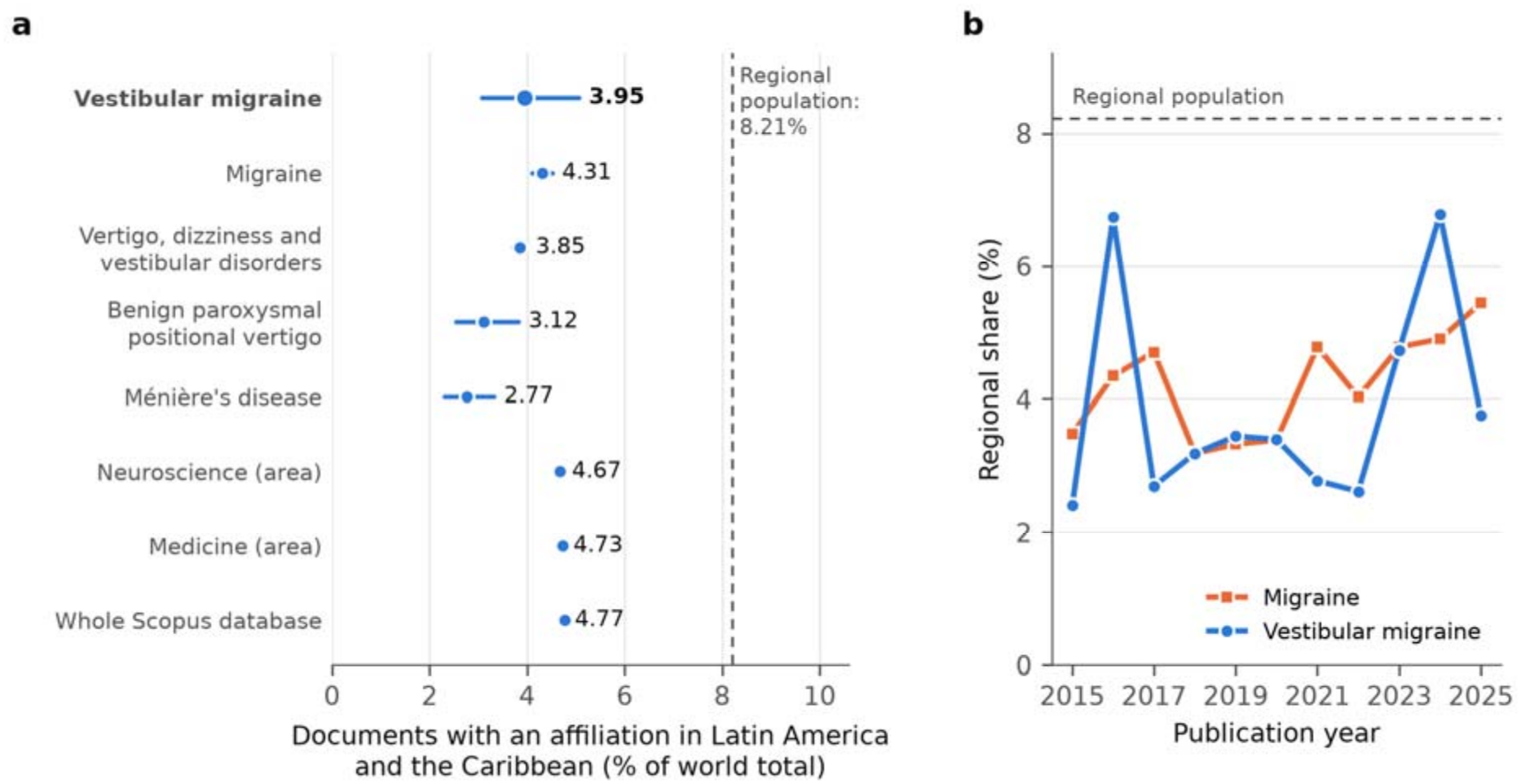
Participation of Latin America and the Caribbean in Scopus-indexed output, 2015-2025. (a) By field, with 95% confidence interval. (b) Annual trend in vestibular migraine and in migraine. The dashed line shows the region’s share of the world population.

### Comparison with other regions

North America produced 1.04 documents per million inhabitants and Europe and Central Asia 0.66, that is, 11.5 and 7.3 times the rate of Latin America and the Caribbean (p < 0.001); East Asia and Pacific produced 2.1 times more (p < 0.001) (Table 1). The region did not differ from the Middle East and North Africa (p = 0.26) and exceeded South Asia and Sub-Saharan Africa (p < 0.001). Europe and Central Asia (index 1.22) and North America (1.17) contributed more to this field than to the whole database; the other regions contributed less, with significant deficits in East Asia and Pacific, Middle East and North Africa, South Asia and Sub-Saharan Africa. The index for Latin America and the Caribbean, 0.83, did not differ from 1 (p = 0.16).

**Table 1.** Vestibular migraine output by world region, Scopus, 2015-2025.

| Region | Documents | % of world total | % of world population | Per million inhabitants | Ratio to Latin America and the Caribbean | % of whole database | Special-ization index (p) |
|---|---|---|---|---|---|---|---|
| North America | 383 | 26.1 | 4.7 | 1.04 | 11.5 | 22.3 | 1.17 (0.001) |
| Europe and Central Asia | 606 | 41.3 | 11.8 | 0.66 | 7.3 | 33.8 | 1.22 (< 0.001) |
| East Asia and Pacific | 442 | 30.1 | 30.1 | 0.19 | 2.1 | 34.5 | 0.87 (< 0.001) |
| Latin America and the Caribbean | 58 | 4.0 | 8.2 | 0.09 | 1.0 | 4.8 | 0.83 (0.16) |
| Middle East and North Africa | 55 | 3.7 | 9.7 | 0.07 | 0.8 | 6.7 | 0.56 (< 0.001) |
| South Asia | 45 | 3.1 | 20.6 | 0.03 | 0.3 | 6.9 | 0.45 (< 0.001) |
| Sub-Saharan Africa | 9 | 0.6 | 14.9 | 0.01 | 0.1 | 2.0 | 0.31 (< 0.001) |

World Bank regions. A document with authors from several regions counts in each. Specialization index: share in vestibular migraine divided by share in the whole database; p from the exact binomial test.

### Distribution within the region and collaboration

Eight of the 34 countries or territories had at least one document: Brazil (38; 59.4%), Chile (10), Colombia (7), Argentina (6), Mexico (4), Guyana (2), Costa Rica (1) and Peru (1) (details in the supplementary material). Thirty-four regional documents (53.1%) included an affiliation from outside the region, compared with 13.9% of multi-country documents in the rest of the corpus (p < 0.001). Only four brought together two or more countries of the region, and all four also included an external institution: none was an exclusively regional collaboration. In the corpus, documents with authors from several countries received more citations (median 10 vs 6; p < 0.001), a difference not observed within the region (9 vs 7.5; p = 0.82), where internationally co-authored documents are more recent (median year 2024 vs 2020).

### Design and orientation

Regional and non-regional documents had the same distribution of designs (chi-squared = 2.44; 6 degrees of freedom; permutation p = 0.88) (Table 2). There was no regional clinical trial, but outside the region there were 24 of 1363 (1.76%), so the absence is compatible with chance (p = 0.62). Therapeutic orientation was less frequent in the region (17.2% vs 27.3%; p = 0.08). The focal subset (p = 0.97) and the PubMed-indexed documents (p = 0.85) gave equivalent results. Median citations were 8 (interquartile range 2 to 16) for regional documents and 6 (1 to 19) for the rest (p = 0.66).

**Table 2.** Design and orientation of documents by regional affiliation.

| **Category** | **Latin America and the Caribbean (n = 64), n (%)** | **Rest of the world (n = 1363), n (%)** | **p** |
|---|---|---|---|
| Randomized controlled trial | 0 (0.0) | 18 (1.3) | 1.00 |
| Other clinical trial or protocol | 0 (0.0) | 6 (0.4) | 1.00 |
| Systematic review or meta-analysis | 2 (3.1) | 55 (4.0) | 1.00 |
| Narrative review | 14 (21.9) | 254 (18.6) | 0.51 |
| Case report or series | 2 (3.1) | 52 (3.8) | 1.00 |
| Observational or other original | 40 (62.5) | 888 (65.2) | 0.69 |
| Letter, editorial or commentary | 6 (9.4) | 90 (6.6) | 0.44 |
| *Therapeutic orientation* | 11 (17.2) | 372 (27.3) | 0.08 |

The first seven rows are mutually exclusive; the last is an independent flag. "Rest of the world" includes 21 documents with undeterminable country. p values from Fisher's exact test.

## Discussion

Our results answer two different questions. Does Latin America and the Caribbean produce less vestibular migraine research than other regions? Yes, by a wide margin: per inhabitant, North America produces more than eleven times and Europe and Central Asia more than seven times what the region produces, and the region contributes 4% of documents with 8% of the world population.

Is that gap larger in this field than in the rest of science? Here we found no evidence: regional participation in vestibular migraine is similar to the region’s participation in migraine, in vestibular disorders and in the database as a whole.

That second answer has a margin. The region’s specialization index is 0.83, a participation 17% below its baseline, and with 58 documents there is no power to distinguish it from 1. We can state that we did not detect an additional lag, not that none exists: a moderate deficit is compatible with what we observed.

Compared with other regions, Latin America and the Caribbean occupies an intermediate position. North America and Europe and Central Asia have a larger share in this field than in the whole database, so the distance from them is larger here (11.5 and 7.3 times per inhabitant) than in science overall (8.1 and 4.9 times). South Asia, the Middle East and North Africa and Sub-Saharan Africa, by contrast, reach between one half and one third of their database share in this field. Outside North America and Europe, Latin America and the Caribbean is the only region whose index does not differ from 1. The predominance of the United States is consistent with other bibliometric studies in the area^10,11^. We did not measure investment, number of researchers or installed capacity, so we describe the magnitude of the gap and not its causes.

In the type of studies, the region does not differ from the rest of the world: in both, observational studies and narrative reviews predominate. The absence of regional clinical trials is not a distinctive feature either: at the frequency observed outside the region, the probability that 64 documents contain none is about one in three, and reviews agree that trial evidence in this condition is scarce and of low quality^4,5^.

Regional output does stand out in three respects. It is concentrated: Brazil participates in six of every ten documents, and 26 of the 34 countries or territories have none. It looks outward: more than half of the documents include external institutions, almost four times the proportion of international collaboration in the rest of the corpus, and none is an exclusively regional collaboration. And it is not growing: the region’s share in migraine increases over the period, but in vestibular migraine it shows no trend.

### Limitations

The comparison between fields was made in a single database. Scopus and PubMed incompletely cover regional journals and those published in Spanish and Portuguese^19^, and we did not search LILACS or SciELO, so regional output is underestimated in absolute terms; the comparison between fields assumes that this underestimation is equal across them. Design classification was automatic and, for documents present only in Scopus, relied on title and document type. Full counting attributes to a country documents in which its contribution may have been minor, and the size of the regional set limits the power to detect moderate differences. The World Bank grouping places Turkey and Central Asia with Europe and Australia with East Asia, and citation counts depend on the date of retrieval.

## Conclusions

Between 2015 and 2025, Latin America and the Caribbean contributed about 4% of the world output on vestibular migraine, half of what its population would suggest. Per inhabitant, North America produced 11.5 times more and Europe and Central Asia 7.3 times more, a distance similar in magnitude to that observed in the whole database. Regional participation in vestibular migraine was comparable to the region’s participation in migraine, in vestibular disorders and in the whole database; we did not detect an additional lag specific to the field, although the size of regional output does not allow a moderate deficit to be ruled out. Nor did we find differences in the type of studies. These conclusions are limited to the literature indexed in Scopus and PubMed and rely on an automatic classification of designs.

The following suggestions are our own proposals based on the findings; the study did not evaluate their effect. Joint studies among the countries of the region that already publish in the field (Brazil, Chile, Colombia, Argentina and Mexico account for almost all regional output and have no exclusively regional collaboration) could be oriented toward designs that are absent or infrequent in the region, such as clinical trials and therapeutically oriented studies, and could incorporate the countries without output through existing groups. The effect of such collaboration should be measured: international collaboration was associated with more citations in the worldwide corpus but not within the region, and this association does not demonstrate causality.

## Supporting information

Additional methodological and analytical details for the bibliometric study on Latin America and the Caribbean (LAC) participation in vestibular migra

## Data Availability

All data produced in the present study are available upon reasonable request to the authors

## Declarations

### Author contributions (CRediT)

CAVD: conceptualization, methodology, supervision, writing – original draft. AAAA: writing – review and editing. DIOT: validation, writing – review and editing. ODOT: data curation, formal analysis, software, visualization, writing – review and editing. All authors approved the final version and consented to its posting as a preprint.

### Ethics statement

The study used only openly available bibliographic metadata (PubMed/MEDLINE, Scopus, World Bank population data); it involved no human participants, samples or personal data, so informed consent was not applicable and approval by an ethics committee was not required. Under article 11 of Resolution 8430 of 1993 of the Colombian Ministry of Health, it is classified as research without risk.

### Data availability

The code for retrieval, cleaning, analysis and figures, the raw database responses and the cleaned dataset are available from the authors upon request to Oscar Daniel Oviedo Tapia. The verbatim search strings, the selection flow, the comparator data and the list of the 64 regional documents are provided in the supplementary material.

### Use of generative artificial intelligence

A large language model (Claude, Anthropic) was used to write the code for retrieval, cleaning, data analysis and figures. The authors reviewed the code, verified the results against the data and the references against their sources, and take full responsibility for the content. No artificial intelligence system is listed as an author.

### Reporting guideline

BIBLIO (Preliminary guideline for reporting bibliometric reviews of the biomedical literature)^15^.

## Acknowledgments

None.

