## Additional methodological and analytical details for the bibliometric study on Latin America and the Caribbean (LAC) participation in vestibular migra for "Participation of Latin America and the Caribbean in vestibular migraine research: a bibliometric study"

Abbreviations: LAC, Latin America and the Caribbean; LA restricted, the 20 Latin American countries plus Puerto Rico (sensitivity definition); VM, vestibular migraine; CI, confidence interval.

### S0. Methodological details

**Record matching and year.** Records were matched first by PubMed identifier, then by DOI and, for records with neither, by normalized title (without accents, punctuation or spaces) and year, accepting only unique matches with titles longer than 25 characters. Records with the same normalized title and first author were considered second versions of the same work; the one indexed in PubMed or, failing that, the most cited was retained. The year assigned was that of first publication: the earliest of the PubMed electronic and print dates and the Scopus cover date.

**Country.** For Scopus records, the search was repeated with the AFFILCOUNTRY filter for each country, and the records returned by each query were recorded, because the standard view of the application programming interface reports only some of the affiliations. For PubMed records, each declared affiliation was resolved with a dictionary of country names, United States states and frequent cities. Both sources were combined by union; when neither provided a country, OpenAlex was used (3 documents).

**Sensitivity.** With the restricted definition (20 Latin American countries and Puerto Rico) the regional set comprised 62 documents (4.34%); with the regional first-author criterion, 47.

### S1. Verbatim search strings

**PubMed/MEDLINE** (searched 2026-10-12; 949 records)

```
("Vestibular Migraine"[Mesh] OR "vestibular migraine"[tiab] OR "vestibular migraine"[ot] OR "vestibular migraines"[tiab] OR "vestibular migraines"[ot] OR "migrainous vertigo"[tiab] OR "migrainous vertigo"[ot] OR "migraine-associated vertigo"[tiab] OR "migraine-associated vertigo"[ot] OR "migraine-related vertigo"[tiab] OR "migraine-related vertigo"[ot] OR "migraine-associated dizziness"[tiab] OR "migraine-associated dizziness"[ot] OR "migraine-related dizziness"[tiab] OR "migraine-related dizziness"[ot] OR "migraine-related vestibulopathy"[tiab] OR "migraine-related vestibulopathy"[ot] OR "vertiginous migraine"[tiab] OR "vertiginous migraine"[ot] OR "migraña vestibular"[tiab] OR "migraña vestibular"[ot] OR "vértigo migrañoso"[tiab] OR "vértigo migrañoso"[ot] OR "migrânea vestibular"[tiab] OR "migrânea vestibular"[ot] OR "enxaqueca vestibular"[tiab] OR "enxaqueca vestibular"[ot]) AND ("2015/01/01"[dp] : "2025/12/31"[dp])
```

Phrases reported by PubMed as not found: "migraine-associated dizziness"[ot]; "vertigo migranoso"[tiab]; "vertigo migranoso"[ot]; "enxaqueca vestibular"[tiab]; "enxaqueca vestibular"[ot].

**Scopus** (searched 2026-10-12; 1468 records)

```
TITLE-ABS-KEY("vestibular migraine" OR "vestibular migraines" OR "migrainous vertigo" OR "migraine-associated vertigo" OR "migraine-related vertigo" OR "migraine-associated dizziness" OR "migraine-related dizziness" OR "migraine-related vestibulopathy" OR "vertiginous migraine" OR "migraña vestibular" OR "vértigo migrañoso" OR "migrânea vestibular" OR "enxaqueca vestibular") AND PUBYEAR > 2014 AND PUBYEAR < 2026
```

### S2. Region

Latin America and the Caribbean (33 states and Puerto Rico): Antigua and Barbuda, Argentina, Bahamas, Barbados, Belize, Bolivia, Brazil, Chile, Colombia, Costa Rica, Cuba, Dominica, Dominican Republic, Ecuador, El Salvador, Grenada, Guatemala, Guyana, Haiti, Honduras, Jamaica, Mexico, Nicaragua, Panama, Paraguay, Peru, Puerto Rico, Saint Kitts and Nevis, Saint Lucia, Saint Vincent and the Grenadines, Suriname, Trinidad and Tobago, Uruguay, Venezuela.

Restricted definition (sensitivity analysis): Argentina, Bolivia, Brazil, Chile, Colombia, Costa Rica, Cuba, Dominican Republic, Ecuador, El Salvador, Guatemala, Haiti, Honduras, Mexico, Nicaragua, Panama, Paraguay, Peru, Puerto Rico, Uruguay, Venezuela.

### S3. Selection flow

| Stage | n |
|---|---|
| PubMed records | 948 |
| Scopus records | 1468 |
| Duplicates within Scopus | 5 |
| Duplicates between databases | 894 |
| — matched by PMID / DOI / title and year | 680 / 213 / 1 |
| Unique records | 1517 |
| Excluded: outside 2015-2025 | 8 |
| Excluded: erratum or retraction notice | 6 |
| Excluded: retracted publication | 0 |
| Excluded: book or book chapter | 67 |
| Excluded: second version of the same work | 9 |
| **Corpus** | **1427** |
| — in both databases / PubMed only / Scopus only | 873 / 53 / 501 |
| With an affiliation in Latin America and the Caribbean | 64 |
| Country not determinable | 21 |

Source of country per document: Scopus + PubMed = 849; Scopus = 500; PubMed = 54; no data = 21; OpenAlex = 3.

Agreement of the regional classification between Scopus and PubMed (n = 849): both yes = 40; Scopus only = 0; PubMed only = 0; both no = 809.

### S4. Comparators in Scopus

| Field | World | LAC | LAC % | 95% CI lower | 95% CI upper | LA restricted | LA restricted % | p vs VM |
|---|---|---|---|---|---|---|---|---|

| | | | | | | | | |
|---|---|---|---|---|---|---|---|---|
| Vestibular migraine | 1468 | 58 | 3.95 | 3.07 | 5.07 | 56 | 3.81 | |
| Migraine | 37214 | 1603 | 4.31 | 4.11 | 4.52 | 1578 | 4.24 | 0.555 |
| Vertigo, dizziness and vestibular disorders | 80362 | 3091 | 3.85 | 3.72 | 3.98 | 3033 | 3.77 | 0.837 |
| Ménière's disease | 4006 | 111 | 2.77 | 2.31 | 3.33 | 110 | 2.75 | 0.028 |
| Benign paroxysmal positional vertigo | 2820 | 88 | 3.12 | 2.54 | 3.83 | 88 | 3.12 | 0.157 |
| Neuroscience (area) | 977864 | 45711 | 4.67 | 4.63 | 4.72 | 45290 | 4.63 | 0.215 |
| Medicine (area) | 10622760 | 502961 | 4.73 | 4.72 | 4.75 | 494713 | 4.66 | 0.176 |
| Whole Scopus database | 42536358 | 2028707 | 4.77 | 4.76 | 4.78 | 2009039 | 4.72 | 0.158 |

Comparator search strings (all with PUBYEAR > 2014 AND PUBYEAR < 2026):

- Migraine: TITLE-ABS-KEY(migraine)

- Vertigo, dizziness and vestibular disorders: TITLE-ABS-KEY(vertigo OR dizziness OR "vestibular disease" *OR "vestibular disorder*")

- Ménière's disease: TITLE-ABS-KEY("meniere" *OR "ménière*")

- Benign paroxysmal positional vertigo: TITLE-ABS-KEY("benign paroxysmal positional vertigo")

- Neuroscience: SUBJAREA(NEUR)

- Medicine: SUBJAREA(MEDI)

- Whole Scopus database: year filter only.

Mean population 2015-2025 (World Bank, SP.POP.TOTL): world = 7839785852; region = 643498330 (8.21%).

### S5. Annual series (Scopus)

| Field | Year | World | LAC | LAC % |
|---|---|---|---|---|
| Vestibular migraine | 2015 | 83 | 2 | 2.41 |
| Vestibular migraine | 2016 | 89 | 6 | 6.74 |
| Vestibular migraine | 2017 | 74 | 2 | 2.7 |
| Vestibular migraine | 2018 | 94 | 3 | 3.19 |
| Vestibular migraine | 2019 | 87 | 3 | 3.45 |
| Vestibular migraine | 2020 | 147 | 5 | 3.4 |
| Vestibular migraine | 2021 | 144 | 4 | 2.78 |
| Vestibular migraine | 2022 | 191 | 5 | 2.62 |
| Vestibular migraine | 2023 | 169 | 8 | 4.73 |
| Vestibular migraine | 2024 | 177 | 12 | 6.78 |
| Vestibular migraine | 2025 | 213 | 8 | 3.76 |
| Migraine | 2015 | 2586 | 90 | 3.48 |
| Migraine | 2016 | 2458 | 107 | 4.35 |
| Migraine | 2017 | 2468 | 116 | 4.7 |
| Migraine | 2018 | 2692 | 86 | 3.19 |
| Migraine | 2019 | 2970 | 99 | 3.33 |

| | | | | |
|---|---|---|---|---|
| Migraine | 2020 | 3480 | 118 | 3.39 |
| Migraine | 2021 | 4000 | 191 | 4.78 |
| Migraine | 2022 | 3971 | 160 | 4.03 |
| Migraine | 2023 | 3974 | 190 | 4.78 |
| Migraine | 2024 | 4207 | 206 | 4.9 |
| Migraine | 2025 | 4408 | 240 | 5.44 |

### S6. Classification rule

Applied in this order to the whole corpus (construir.py, function clasificar):

1. Letter, editorial or commentary: PubMed publication type *Letter*, *Editorial*, *Comment* or *News*; in the absence of PubMed types, Scopus type *Letter*, *Editorial* or *Note*.
2. Systematic review or meta-analysis: the corresponding PubMed type, or the expressions "systematic review", "meta-analysis", "scoping review" or "umbrella review" in the title.
3. Randomized controlled trial: PubMed type *Randomized Controlled Trial*, or "randomized", "placebo" or "double-blind" in the title of a document that is neither a review nor a protocol.
4. Other clinical trial or protocol: other PubMed *Clinical Trial* types.
5. Narrative review: PubMed type *Review*, or Scopus *Review* or *Short Survey*.
6. Case report or series: PubMed type *Case Reports* or an equivalent expression in the title.
7. Observational or other original: the rest.

Therapeutic orientation: treatment terms in the title or author keywords, or MeSH qualifiers *drug therapy*, *therapy*, *therapeutic use*, *rehabilitation* or *prevention & control*. Focal subset: the condition is named in the title or author keywords.

**Design and orientation, whole corpus** (region n = 64; rest n = 1363; permutation p = 0.881)

| Category | LAC n | LAC % | Rest n | Rest % | Fisher p |
|---|---|---|---|---|---|
| randomized controlled trial | 0 | 0.0 | 18 | 1.32 | 1.000 |
| other clinical trial/protocol | 0 | 0.0 | 6 | 0.44 | 1.000 |
| systematic review/meta-analysis | 2 | 3.12 | 55 | 4.04 | 1.000 |
| narrative review | 14 | 21.88 | 254 | 18.64 | 0.513 |
| case report/series | 2 | 3.12 | 52 | 3.82 | 1.000 |
| observational/other original | 40 | 62.5 | 888 | 65.15 | 0.688 |
| letter/editorial/commentary | 6 | 9.38 | 90 | 6.6 | 0.437 |
| any clinical trial | 0 | 0.0 | 24 | 1.76 | 0.622 |
| evidence synthesis or trial | 2 | 3.12 | 79 | 5.8 | 0.578 |

| | | | | | |
|---|---|---|---|---|---|
| therapeutic topic | 11 | 17.19 | 372 | 27.29 | 0.083 |
| mentions CGRP or antibodies/gepants | 3 | 4.69 | 52 | 3.82 | 0.734 |

**Design and orientation, focal subset** (region n = 32; rest n = 731; permutation p = 0.973)

| Category | LAC n | LAC % | Rest n | Rest % | Fisher p |
|---|---|---|---|---|---|
| randomized controlled trial | 0 | 0.0 | 10 | 1.37 | 1.000 |
| other clinical trial/protocol | 0 | 0.0 | 4 | 0.55 | 1.000 |
| systematic review/meta-analysis | 1 | 3.12 | 28 | 3.83 | 1.000 |
| narrative review | 7 | 21.88 | 129 | 17.65 | 0.487 |
| case report/series | 2 | 6.25 | 34 | 4.65 | 0.659 |
| observational/other original | 21 | 65.62 | 490 | 67.03 | 0.850 |
| letter/editorial/commentary | 1 | 3.12 | 36 | 4.92 | 1.000 |
| any clinical trial | 0 | 0.0 | 14 | 1.92 | 1.000 |
| evidence synthesis or trial | 1 | 3.12 | 42 | 5.75 | 1.000 |
| therapeutic topic | 6 | 18.75 | 229 | 31.33 | 0.171 |
| mentions CGRP or antibodies/gepants | 2 | 6.25 | 41 | 5.61 | 0.700 |

**Design and orientation, documents retrieved from PubMed** (region n = 47; rest n = 879; permutation p = 0.908)

| Category | LAC n | LAC % | Rest n | Rest % | Fisher p |
|---|---|---|---|---|---|
| randomized controlled trial | 0 | 0.0 | 10 | 1.14 | 1.000 |
| other clinical trial/protocol | 0 | 0.0 | 6 | 0.68 | 1.000 |
| systematic review/meta-analysis | 1 | 2.13 | 40 | 4.55 | 0.717 |
| narrative review | 11 | 23.4 | 169 | 19.23 | 0.453 |
| case report/series | 2 | 4.26 | 37 | 4.21 | 1.000 |
| observational/other original | 32 | 68.09 | 583 | 66.33 | 0.875 |
| letter/editorial/commentary | 1 | 2.13 | 34 | 3.87 | 1.000 |
| any clinical trial | 0 | 0.0 | 16 | 1.82 | 1.000 |
| evidence synthesis or trial | 1 | 2.13 | 56 | 6.37 | 0.355 |
| therapeutic topic | 9 | 19.15 | 264 | 30.03 | 0.139 |
| mentions CGRP or antibodies/gepants | 2 | 4.26 | 42 | 4.78 | 1.000 |

### S7. Documents with an affiliation in Latin America and the Caribbean (n = 64)

Automatic classification. Focal: 1 if the condition is named in the title or author keywords. Cites: Scopus citations at retrieval (blank if not indexed in Scopus). Countries as ISO 3166-1 alpha-2 codes.

| ID | Year | Title | Journal | Countri | Design | Foca | Cite | DOI |
|---|---|---|---|---|---|---|---|---|

| | | | | es | | l | s | |
|---|---|---|---|---|---|---|---|---|
| A1-0898 | 2015 | Vestibular migraine: clinical and epidemiological aspects. | Braz J Otorhinolaryngol | BR | observational/other original | 1 | 43 | 10.1016/j.bjorl.2015.06.003 |
| A1-0906 | 2015 | Vestibular migraine: comparative analysis between diagnostic criteria. | Braz J Otorhinolaryngol | BR | observational/other original | 1 | 10 | 10.1016/j.bjorl.2015.07.007 |
| A1-0864 | 2016 | Balance disorders in childhood: Main etiologies according to age. Usefulness of the video head impulse test. | Int J Pediatr Otorhinolaryngol | AR | observational/other original | 0 | 44 | 10.1016/j.ijporl.2016.06.020 |
| A1-0835 | 2016 | Commentary: Distribution of 5-HT1F Receptors in Monkey Vestibular and Trigeminal Ganglion Cells. | Front Neurol | BR | letter/editorial/commentary | 1 | 1 | 10.3389/fneur.2016.00234 |
| A1-0866 | 2016 | Prophylactic treatment of vestibular migraine. | Braz J Otorhinolaryngol | BR | observational/other original | 1 | 42 | 10.1016/j.bjorl.2016.04.022 |
| A1-1083 | 2016 | Spectrum of migraine variants and beyond: The individual syndromes in children | Brain and Development | GY; IN; US | narrative review | 0 | 9 | 10.1016/j.braindev.2015.05.009 |
| A1-0889 | 2016 | Vestibular Migraine: Vestibular Symptom May Identify Different Subgroups. | Otol Neurotol | BR | observational/other original | 1 | 5 | 10.1097/mao.0000000000000956 |
| A1-0872 | 2016 | Vestibular migraine: diagnosis challenges and need for targeted treatment. | Arq Neuropsiquiatr | BR | narrative review | 1 | 21 | 10.1590/0004-282x20160037 |
| A1-0786 | 2017 | Body balance at static posturography in vestibular migraine. | Braz J Otorhinolaryngol | BR | observational/other original | 1 | 37 | 10.1016/j.bjorl.2017.12.001 |
| A1-1358 | 2017 | Vertebral artery dissection: an important differential diagnosis of vertigo | Brazilian Journal of Otorhinolaryngology | BR | observational/other original | 0 | 0 | 10.1016/j.bjorl.2015.08.020 |
| A1-1211 | 2018 | Episodic status migrainosus: One more case | Cephalalgia | BR | letter/editorial/commentary | 0 | 2 | 10.1177/0333102417710021 |
| A1-0737 | 2018 | Vestibular Migraine with Visual Aura and Olfactory Hallucination in Children: Two Case Reports. | Neuropediatrics | BR | case report/series | 1 | 4 | 10.1055/s-0038-1673642 |
| A1-0731 | 2018 | [Approach to patients with vertigo]. | Medicina (B Aires) | AR | observational/other original | 1 | 7 | |
| A1-0720 | 2019 | Abnormal visuo-vestibular interactions in vestibular migraine: a cross sectional study. | Brain | BR; CH; GB | observational/other original | 1 | 83 | 10.1093/brain/awy355 |
| A1-0697 | 2019 | Auditory brainstem function in women with vestibular migraine: a controlled study. | BMC Neurol | BR; CA | observational/other original | 1 | 15 | 10.1186/s12883-019-1368-5 |
| A1-0665 | 2019 | Physical therapy and migraine: musculoskeletal and balance dysfunctions and their relevance for clinical practice. | Braz J Phys Ther | BR; DE; PL | narrative review | 1 | 73 | 10.1016/j.bjpt.2019.11.001 |
| A1-0683 | 2019 | Postural control at posturography with virtual reality in the intercritical period of vestibular migraine. | Braz J Otorhinolaryngol | BR | observational/other original | 1 | 9 | 10.1016/j.bjorl.2019.06.015 |
| A1-0992 | 2020 | Classification of Tinnitus: Multiple Causes with the | Otolaryngologic Clinics of | BR; US | narrative review | 0 | 27 | 10.1016/j.otc.2020.03.015 |

| | | | | | | | | |
|---|---|---|---|---|---|---|---|---|
| | | Same Name | North America | | | | | |
| A1-0623 | 2020 | Subdiagnosis, but not presence of vestibular symptoms, predicts balance impairment in migraine patients - a cross sectional study. | J Headache Pain | BR; DE; IT | observational/other original | 1 | 14 | 10.1186/s10194-020-01128-z |
| A1-0612 | 2020 | Subjective Visual Vertical Testing in Patients With Chronic Suppurative Otitis Media. | Otolaryngol Head Neck Surg | BR | observational/other original | 0 | 6 | 10.1177/01945998209314 91 |
| A1-0581 | 2020 | Vestibular Syndromes in Childhood and Adolescence. | Int Arch Otorhinolaryngol | BR | observational/other original | 0 | 12 | 10.1055/s-0039-3402443 |
| A1-0591 | 2020 | Vestibular and balance disorders in Mexican children and adolescents: Review of 8-year clinical records. | Acta Otorrinolaringol Esp (Engl Ed) | MX | observational/other original | 0 | 1 | 10.1016/j.otorri.2020.02.011 |
| A1-0644 | 2020 | Video head impulse test in vestibular migraine. | Braz J Otorhinolaryngol | BR | observational/other original | 1 | 23 | 10.1016/j.bjorl.2019.12.009 |
| A1-1008 | 2021 | Does the video head impulse test replace caloric testing in the assessment of patients with chronic dizziness? A systematic review and meta-analysis | Brazilian Journal of Otorhinolaryngology | BR | systematic review/meta-analysis | 0 | 23 | 10.1016/j.bjorl.2021.01.002 |
| A1-0505 | 2021 | Vestibular Migraine and Tinnitus: A Challenging Narrative. | Cureus | MX; US | narrative review | 1 | | 10.7759/cureus.15998 |
| A1-0434 | 2022 | Atypical Positional Vertigo: Definition, Causes, and Mechanisms. | Audiol Res | AR; PT | narrative review | 1 | 16 | 10.3390/audiolres12020018 |
| A1-0374 | 2022 | Caloric and video head impulse test dissociated results in dizzy patients. | Front Neurol | CL | observational/other original | 0 | 12 | 10.3389/fneur.2022.1000318 |
| A1-0392 | 2022 | Dissociation between Caloric and Video Head Impulse Tests in Dizziness Clinics. | Audiol Res | CL | narrative review | 1 | 16 | 10.3390/audiolres12040043 |
| A1-0379 | 2022 | Facts of Vertigo in Adolescents: Controversies and Challenges - A Narrative Review. | Cureus | CO; MX; US | narrative review | 1 | | 10.7759/cureus.28294 |
| A1-0371 | 2022 | Head shaking does not alter vestibulo ocular reflex gain in vestibular migraine. | Front Neurol | AR; CL; GB | observational/other original | 1 | 6 | 10.3389/fneur.2022.967521 |
| A1-0420 | 2022 | Migraine and balance impairment: Influence of subdiagnosis, otoneurological function, falls, and psychosocial factors. | Headache | BR; CA; DE; PL | observational/other original | 1 | 23 | 10.1111/head.14309 |
| A1-0410 | 2022 | Vertigo in Pregnancy: A Narrative Review. | Cureus | CO; US | narrative review | 0 | | 10.7759/cureus.25386 |
| A1-0396 | 2022 | Vestibular migraine. | Arq Neuropsiquiatr | BR | observational/other original | 1 | 9 | 10.1590/0004-282x-anp-2022-s111 |
| A1-0435 | 2022 | [Osmophobia and vestibular migraine]. | Rev Fac Cien Med Univ Nac Cordoba | CL; US | case report/series | 1 | | 10.31053/1853.0605.v79.n1.33699 |
| A1-1381 | 2023 | A new treatment for cervical dizziness | Brazilian Journal of Otorhinolaryngology | BR | observational/other original | 0 | 0 | 10.1016/j.bjorl.2023.101321 |
| A1-1010 | 2023 | Acute truncal ataxia without nystagmus in patients with | European Journal of | AR; GB | observational/other | 0 | 22 | 10.1111/ene.15729 |

| | | | | | | | | |
|---|---|---|---|---|---|---|---|---|
| | | acute vertigo | Neurology | | original | | | |
| A1-0307 | 2023 | Balance disorders and migraine. | Musculoskelet Sci Pract | BR; DE | observational/other original | 0 | 13 | 10.1016/j.msksp.2023.102783 |
| A1-0344 | 2023 | Cross-cultural adaptation of the Dizziness Handicap Inventory - child/adolescent to Spanish version. Analysis of its application in children. | Acta Otorrinolaringol Esp (Engl Ed) | AR | observational/other original | 0 | 2 | 10.1016/j.otoeng.2023.01.002 |
| A1-1389 | 2023 | Editorial: Audiovestibular disorders in pediatrics | Frontiers in Pediatrics | CL; IT; US | letter/editorial/commentary | 0 | 0 | 10.3389/fped.2023.1231803 |
| A1-1107 | 2023 | Migraine Attacks Triggered by Ingestion of Watermelon | European Neurology | BR | observational/other original | 0 | 7 | 10.1159/000531286 |
| A1-0279 | 2023 | Vestibular and Oculomotor Findings in Vestibular Migraine Patients. | Audiol Res | CL | observational/other original | 1 | 15 | 10.3390/audiolres13040053 |
| A1-1261 | 2024 | Brazilian Version of the Vestibular Activities and Participation Measure: Cross-Cultural Adaptation, Validity, and Reliability | Journal of Otolaryngology Head and Neck Surgery | BR; US | observational/other original | 0 | 1 | 10.1177/19160216241250353 |
| A1-0180 | 2024 | Clinical Profile, Trends, and Management in Pediatric Patients with Audiovestibular Disorders: Can We Predict Emotional Disability in Pediatric Patients with Episodes of Vertigo and Dizziness? | Audiol Res | CL; ES | observational/other original | 0 | 3 | 10.3390/audiolres14040059 |
| A1-1294 | 2024 | Editorial: Benign paroxysmal positional vertigo | Frontiers in Neurology | BR; ES; IN | letter/editorial/commentary | 0 | 1 | 10.3389/fneur.2024.1358860 |
| A1-0170 | 2024 | Evaluation of cervical vestibular miogenic evoked potential and electrococleography in the diagnosis of vestibular migraine. | Braz J Otorhinolaryngol | BR | observational/other original | 1 | 0 | 10.1016/j.bjorl.2024.101489 |
| A1-1094 | 2024 | Functional Dizziness as a Spatial Cognitive Dysfunction | Brain Sciences | CL | observational/other original | 0 | 8 | 10.3390/brainsci14010016 |
| A1-0161 | 2024 | Imaging of Vertigo and Dizziness: A Site-based Approach Part 3 (Brainstem, Cerebellum, and Miscellaneous). | Semin Ultrasound CT MR | BR | narrative review | 0 | 0 | 10.1053/j.sult.2024.09.005 |
| A1-1303 | 2024 | Increased frequency and intensity of complicated migraine sans migraine after third BNT162b2 dose | Clinics | AT; BR | letter/editorial/commentary | 0 | 1 | 10.1016/j.clinsp.2023.100315 |
| A1-0178 | 2024 | Motion sickness whilst reading as a passenger in the car is highly predictive of vestibular migraine. | Front Neurol | BR; GB; GR | observational/other original | 1 | 4 | 10.3389/fneur.2024.1426081 |
| A1-0958 | 2024 | Redefining migraine prevention: Early treatment with anti-CGRP monoclonal antibodies enhances response in the real world | Journal of Neurology Neurosurgery and Psychiatry | CO; DE; ES; GB; IT; NO; PL; PT; SE | observational/other original | 0 | 66 | 10.1136/jnnp-2023-333295 |
| A1-0230 | 2024 | Sense of direction in vestibular disorders. | J Vestib Res | CL; GB | observational/other original | 0 | 4 | 10.3233/ves-230082 |
| A1-1204 | 2024 | Vestibular Disorders Limited | International | BR | observationa | 0 | 2 | 10.1055/s- |

| | | | | | | | | |
|---|---|---|---|---|---|---|---|---|
| | | to the Vertical Semicircular Canals | Archives of Otorhinolaryngology | | observational/other original | | | 0044-1786047 |
| A1-1243 | 2024 | Vestibular drop attacks diagnosed with valuable insights from CCTV | Lancet | AM; BR; GB | letter/editorial/commentary | 0 | 1 | 10.1016/s0140-6736(24)01589-7 |
| A1-0033 | 2025 | Evaluating associations of migraine-related vertigo. | Headache | BE; GB; PE; TH | observational/other original | 1 | | 10.1111/head.15038 |
| A1-0106 | 2025 | Exploring the Link Between Traumatic Brain Injury and Benign Paroxysmal Positional Vertigo. | Cureus | BR; CO; MX; US | narrative review | 0 | | 10.7759/cureus.81847 |
| A1-0067 | 2025 | Insights into Vestibular Migraine: Diagnostic Challenges, Differential Spectrum and Therapeutic Horizons. | J Clin Med | CO; ES; GB | narrative review | 1 | 12 | 10.3390/jcm14144828 |
| A1-0071 | 2025 | Integrated diagnostic algorithm for acute vertigo combining TiTrATE, STANDING, and HINTS: a validation study in the emergency department. | Sci Rep | CA; CL; GB | observational/other original | 0 | 9 | 10.1038/s41598-025-11007-9 |
| A1-0134 | 2025 | Is there a difference in the results of the video head impulse test in patients with a nosological diagnosis of Ménière's Disease and Vestibular Migraine? | Codas | BR | observational/other original | 1 | 2 | 10.1590/2317-1782/e20230359pt |
| A1-0028 | 2025 | Prophylactic Management of Vestibular Migraine: A Systematic Review. | Ann Clin Transl Neurol | GY; SA | systematic review/meta-analysis | 1 | 3 | 10.1002/acn3.70234 |
| A1-0126 | 2025 | Self-reported physical activity differentiates vestibular migraine from persistent postural perceptual dizziness. | J Vestib Res | BR; GB | observational/other original | 1 | 3 | 10.1177/09574271251318971 |
| A1-0121 | 2025 | Sex differences in dizziness diagnoses across acute and chronic neurological settings. | Neurol Sci | BR; GB; IT | observational/other original | 0 | 11 | 10.1007/s10072-025-08085-y |
| A1-1463 | 2025 | The Impact of Neuroglia on Vestibular Disorders: Insights and Implications | Neuroglia | CO; CR; US | narrative review | 0 | 0 | 10.3390/neuroglia6010010 |
| A1-0130 | 2025 | The Near Point of Convergence in Patients with Vestibular Migraine. | Int Arch Otorhinolaryngol | BR; PT | observational/other original | 1 | 2 | 10.1055/s-0044-1791643 |
| A1-0063 | 2025 | Therapeutic Management of Vestibular Disorders During Pregnancy: A Narrative and Evidence-Based Review. | Cureus | CO; DK; IN; TR; US | narrative review | 0 | | 10.7759/cureus.89705 |

### S8. Documents with an affiliation in Latin America and the Caribbean, by country

| Country | Documents, n (% of 64) | First author from the country | With co-authors from another country | Scopus citations | Documents per million inhabitants |
|---|---|---|---|---|---|
| Brazil | 38 (59.4) | 31 | 16 | 530 | 0.18 |
| Chile | 10 (15.6) | 5 | 6 | 73 | 0.52 |
| Colombia | 7 (10.9) | 6 | 7 | 78 | 0.14 |
| Argentina | 6 (9.4) | 4 | 3 | 97 | 0.13 |

| | | | | | |
|---|---|---|---|---|---|
| Mexico | 4 (6.2) | 1 | 3 | 1 | 0.03 |
| Guyana | 2 (3.1) | 0 | 2 | 12 | 2.49 |
| Costa Rica | 1 (1.6) | 0 | 1 | 0 | 0.20 |
| Peru | 1 (1.6) | 0 | 1 | 0 | 0.03 |

Full counting: a document with authors from several countries counts in each, so the sum exceeds 64. The remaining 26 countries or territories had no documents.
